# COVID-19 outbreak at a large homeless shelter in Boston: Implications for universal testing

**DOI:** 10.1101/2020.04.12.20059618

**Authors:** Travis P. Baggett, Harrison Keyes, Nora Sporn, Jessie M. Gaeta

## Abstract

The circumstances of homelessness create the potential for rapid transmission of SARS-CoV-2 in this vulnerable population. Upon observing a cluster of COVID-19 cases from a single large homeless shelter in Boston, Boston Health Care for the Homeless Program conducted symptom assessments and polymerase chain reaction (PCR) testing for SARS-CoV-2 among all guests residing at the shelter over a 2-day period. Of 408 participants, 147 (36.0%) were PCR-positive for SARS-CoV-2. COVID-positive individuals were more likely to be male (p<0.001) but did not differ significantly from COVID-negative individuals with respect to other demographic and clinical characteristics. Cough (7.5%), shortness of breath (1.4%), and fever (0.7%) were all uncommon among COVID-positive individuals. Our findings illustrate the rapidity with which COVID-19 can be widely transmitted in a homeless shelter setting and suggest that universal PCR testing, rather than a symptom triggered approach, may be a better strategy for identifying and mitigating COVID-19 among people experiencing homelessness.

## Introduction

In the US, an estimated 2.3-3.5 million people experience homelessness annually,^1^ and about 500,000 individuals sleep in homeless shelters each night.^2^ The congregate nature and hygienic challenges of shelter life create the potential for rapid transmission of SARS-CoV-2 in this vulnerable population.

In mid-March, 2020, Boston Health Care for the Homeless Program (BHCHP), in partnership with city and state public health agencies and community partners, created a COVID-19 response strategy that included front-door symptom screening at area shelters, expedited referrals for SARS-CoV-2 testing and isolation for those with respiratory symptoms, dedicated treatment settings for COVID-positive individuals, and detailed contact tracing of confirmed cases.

During the last week in March, BHCHP observed an evolving cluster of COVID-19 cases emerging from a single large homeless shelter in Boston. With support from the Massachusetts Department of Public Health (MDPH), BHCHP rapidly conducted polymerase chain reaction (PCR) testing for SARS-CoV-2 along with focused symptom assessments among all guests residing at the shelter over a 2-day period. In this report, we describe the proportion of tests returning positive and the symptom profile of confirmed cases.

## Methods

Participants were adults ≥18 years old residing in a large homeless shelter in Boston during the onset of the COVID-19 case cluster described above. The original cases (N=15) were identified sequentially over a 5-day period, and each was expeditiously removed from the shelter population at the time of symptom recognition. These individuals predated the implementation of universal testing procedures and are excluded from this study.

Among all remaining shelter guests, BHCHP staff assessed self-reported age, gender, race, and ethnicity. Participants were asked about cough and shortness of breath and given the option to report other symptoms. Body temperature measurements were obtained using oral thermometers. Nasopharyngeal specimens were collected by BHCHP clinical staff using a polyester swab and sent to the MDPH State Public Health Laboratory for SARS-CoV-2 PCR testing.

We used descriptive statistics to characterize the study sample, the proportion of positive PCR tests, and the symptom profile of confirmed cases. We used t tests and Fisher exact tests to compare the characteristics of COVID-positive and COVID-negative individuals.

This study was exempted by the Partners HealthCare Human Research Committee with a waiver of informed consent.

## Results

A total of 408 individuals underwent symptom assessment and COVID-19 PCR testing. The mean age was 51.6 years, 71.6% were male, 33.1% were Black or African American, and 18.6% were Hispanic or Latino (Table). Among all participants, 8.1% reported cough, 0.7% reported shortness of breath, and 5.9% reported other symptoms, including 1.2% with diarrhea. Mean body temperature was 98.4◻.

**Table 1.**
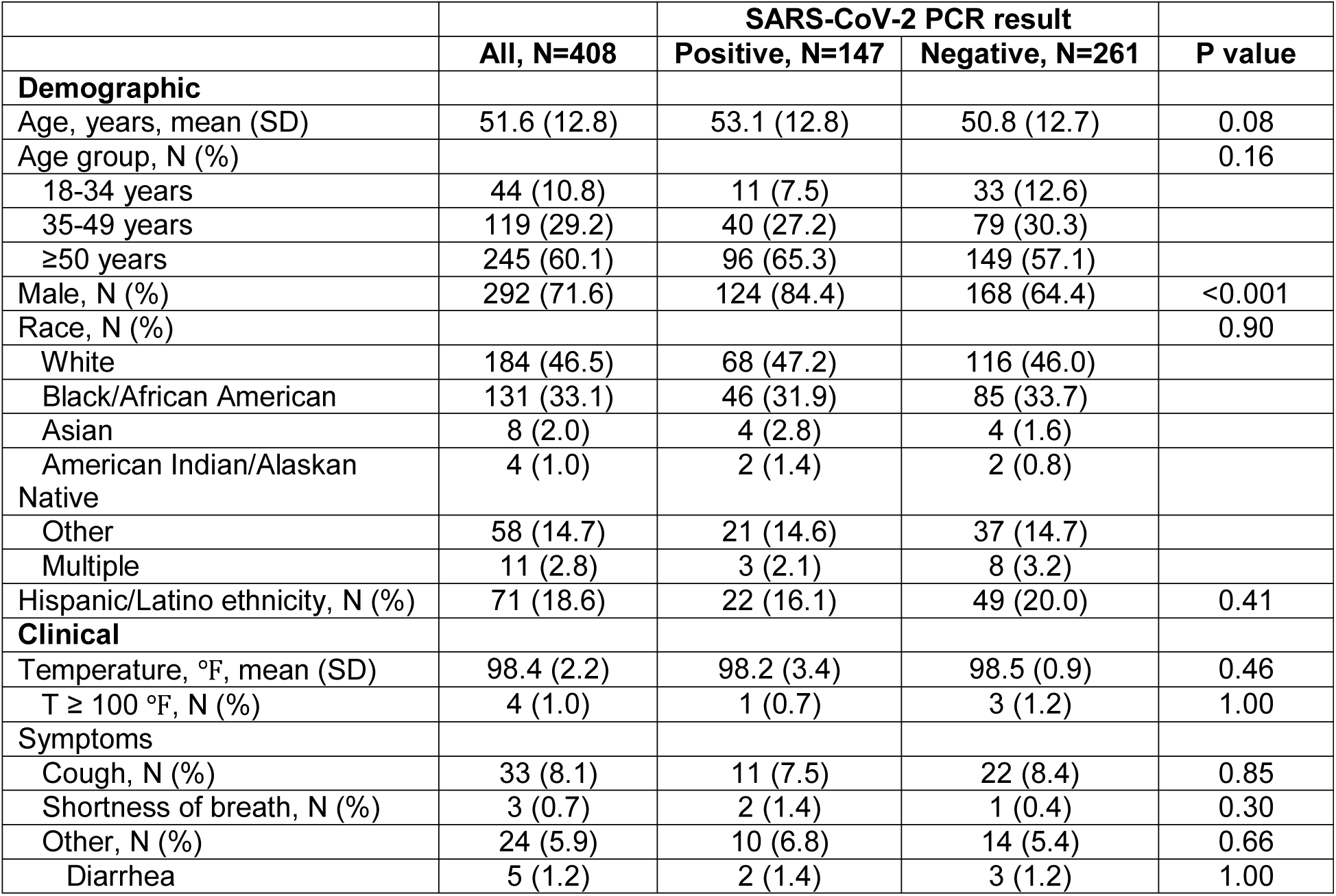
Characteristics of the study sample, overall and by SARS-CoV-2 PCR result.

One hundred forty-seven participants (36.0%) were PCR-positive for SARS-CoV-2. COVID-positive individuals were more likely to be male (p<0.001) but did not differ significantly from COVID-negative individuals with respect to other demographic or clinical characteristics. Cough (7.5%), shortness of breath (1.4%), and fever (0.7%) were all uncommon among COVID-positive individuals.

## Discussion

Universal SARS-CoV-2 PCR testing of an adult homeless shelter population in Boston shortly after the identification of a COVID-19 case cluster yielded an alarming 36% positivity rate. The vast majority of newly identified cases had no symptoms and no fever on a single point-in-time assessment. Our findings illustrate the rapidity with which COVID-19 can be widely transmitted within a homeless shelter setting, even when infection control vigilance is high. Although recommended by the Centers for Disease Control and Prevention^3^ and widely implemented in Boston and elsewhere, front-door symptom screening in homeless shelter settings will likely miss a substantial number of COVID-19 cases in this high-risk population. These results support a universal PCR testing approach for identifying COVID-19 among people experiencing homelessness.

## Data Availability

The study data are not available to share due to the highly vulnerable nature of the patient population.

## Acknowledgments

We thank the Massachusetts Department of Public Health for facilitating the testing described in this paper. We thank Joana Barbosa Teixeira, Andrea Joyce, Elijah Rodriguez, and Erin Ford at Massachusetts General Hospital, and Alexei Alvarado at Boston Health Care for the Homeless Program, for their assistance with data entry. We thank Boston Health Care for the Homeless Program staff for their dedication to patient care. We thank homeless shelter staff for the work they do each day.

Dr. Baggett receives royalties from UpToDate for authorship of a topic review on homeless health care.

## Notes

### Funding Statement

No external funding supported this study.

